# CLINICAL TRAJECTORY AND GENETIC LANDSCAPE OF NEONATAL-ONSET HYPERTROPHIC CARDIOMYOPATHY: INSIGHTS FROM A 30-YEAR MULTICENTER COHORT STUDY

**DOI:** 10.64898/2026.07.06.26357420

**Authors:** Rachele Adorisio, Nicoletta Cantarutti, Sara Di Marzio, Gessica Ingrasciotta, Alessio Franceschini, Elena Cavarretta, Carolina D’Anna, Erica Mencarelli, Diego Martinelli, Massimo Stefano Silvetti, Fabrizio Drago, Cosimo Marco Campanale, Marco Masci, Antonio Novelli, Monia Magliozzi, Luca Di Chiara, Lorenzo Galletti, Flaminia Calzolari, Irma Capolupo, Antonio Amodeo, Andrea Dotta, Alessandra Toscano, Cooperating Investigators

## Abstract

**Background:** Neonatal-onset hypertrophic cardiomyopathy (HCM) is a rare condition with limited data regarding clinical presentation, genetic background, and long-term outcomes. We aimed to characterize the phenotype and prognosis of HCM presenting in neonates.

**Methods:** This is a multicenter retrospective study including patients diagnosed with HCM before 1 year of age. Neonatal-onset HCM was defined as presentation ≤28 days of life. Clinical, genetic, instrumental data, treatment, and outcomes were collected. Primary outcome included overall and cardiac survival, major arrhythmic events (MAEs), implantable cardioverter-defibrillator (ICD) implantation, and cardiac surgery.

**Results:** Among 321 pediatric HCM, 21% were diagnosed during infancy and 75% were neonates. Median age at diagnosis was 1 day (IQR 0–6), 82% presented within the first week of life. Prenatal suspicion was in 25%. At presentation, 41% were symptomatic. RASopathies represented the most common etiology (41%), followed by gene-elusive (31%), mitochondrial/inborn errors of metabolism (18%), and sarcomeric (8%). Left ventricular outflow tract obstruction was frequent in sarcomeric and RASopathy. Overall survival was 92% and cardiac survival was 96% at 2 years; long-term survival was 88% at 30 years. ICDs were implanted in 8%; 21% required cardiac surgery. Survival free from ICD was 40% at 15 year and 47% from myectomy. All events occurred in patients presenting within the first weeks of life.

**Conclusions:** Neonatal-onset HCM is characterized by etiologic heterogeneity, predominance of syndromic and non-sarcomeric etiologies, and long-term cardiovascular morbidity. Presentation within the first days of life identifies a high-risk subgroup requiring intensive surveillance and specialized multidisciplinary management.

## INTRODUCTION

Neonatal hypertrophic cardiomyopathy (HCM) represents a rare but clinically challenging form of pediatric cardiomyopathy, characterized by marked etiologic heterogeneity, variable clinical presentation, and an incompletely defined natural history [1–4]. Unlike HCM presenting during later childhood or adulthood [5–8], neonatal-onset disease is frequently associated with syndromic, metabolic, or mitochondrial disorders rather than isolated sarcomeric mutations, and often manifests with severe heart failure (HF), respiratory compromise, or hemodynamic instability within the first weeks of life. Despite major advances in prenatal diagnosis, molecular genetics, and pediatric HF management, data specifically focused on neonatal HCM remain scarce and are largely limited to small single-center series or heterogeneous infant cohorts [2–7].

Early-onset HCM is increasingly recognized during fetal life or immediately after birth owing to the widespread use of fetal echocardiography and early neonatal cardiovascular screening. However, the prognostic implications of myocardial hypertrophy detected during the neonatal period remain poorly understood. Previous studies have suggested that younger age at presentation may be associated with worse outcomes [1, 5–11], including progressive HF, arrhythmic events, and need for heart transplantation [12], but the extent to which neonatal presentation identifies a biologically distinct and particularly aggressive disease phenotype remains uncertain. In addition, the relative contribution of different genetic etiologies to phenotype severity and long-term outcomes in neonates has not been comprehensively characterized.

Neonatal HCM also poses important diagnostic and therapeutic challenges. Because of the rarity and marked etiology and biological heterogeneity of neonatal HCM evidence supporting medical treatment, risk stratification and timing of advanced therapies in this age group is extremely limited [2,3, 13–15]. As survival improves, understanding the long-term evolution of neonatal HCM has become increasingly relevant for counseling, surveillance, and management planning.

Therefore, the present multicenter study aimed to characterize the demographic, clinical presentation, genetic background, echocardiographic assessment of neonatal HCM, including short- and long-term outcomes in terms of overall and cardiac mortality and major cardiovascular events. In particular, we sought to evaluate the impact of timing of presentation on disease severity and prognosis, and to identify the clinical features associated with adverse cardiovascular outcomes in this uniquely vulnerable population.

## METHODS

### Study Design

This is a multi-center retrospective cohort study including all consecutive neonates and infants (<1^st^ year) presenting between January 1992 and August 2025 with a definitive diagnosis of HCM and referred to Bambino Gesù Children’s Hospital, IRCCS as tertiary center, as well as from other participating pediatric hospitals across Italy. HCM was defined as ventricular thickness >2 z-score in at least one segment on echocardiography, in absence of secondary causes of pathological ventricular hypertrophy. According to the American Heart Association definition for pediatric HCM [1], secondary causes of pathological hypertrophy, including systemic hypertension, aortic stenosis, congenital heart disease, endocrine disorders (e.g., infants of diabetic mothers) or metabolic conditions such as Pompe disease receiving enzyme replacement therapy, were excluded.

Patients were categorized in 2 groups based on the age at first evaluation, as:

- Age ≤28 days at presentation for neonatal HCM;
- Age >28 days but <1 year at presentation for infantile HCM;

Demographic characteristics, prenatal diagnosis, clinical presentation, symptoms, genetic findings, electrocardiographic (ECG) and echocardiographic parameters, medical management, and clinical outcomes were systematically collected. Overall and cardiac survival were defined as freedom from death from any cause and freedom from cardiac death or heart transplantation, respectively. Data were collected at baseline, at 2-year follow-up (FU), and at last available FU. Clinical outcomes included also: 1) left and/or right ventricular outflow tract (LVOT and RVOT respectively) obstruction requiring surgical intervention; 2) Ventricular assist device (VAD) implantation; 3) composite endpoint of major arrhythmic events (MAEs), including sudden cardiac death, aborted sudden cardiac death, and appropriate implantable cardioverter-defibrillator shock (ICD); 4) ICD implantation in primary prevention.

Clinical status at presentation was assessed using age-appropriate parameters to evaluate HF severity, including Ross classification [16] as well as growth parameters (weight and length percentiles).

Patients admitted for acute decompensated HF were additionally stratified according to stage [17]. Acute decompensated HF was defined by the presence of at least one of the following: oxygen saturation <92%, lactate > 2 mmol/L, need for respiratory support, signs of respiratory distress (tachypnea, retractions), or hemodynamic instability [18].

Echocardiographic assessment was performed according to current pediatric echocardiography recommendations [19]. Left ventricular wall thickness and chamber dimensions were measured and expressed as z-scores when appropriate. Parameters analyzed included systolic function, left and right ventricular outflow tract obstruction. LV mass was calculated using standard echocardiographic methods and indexed to body size using pediatric-specific allometric scaling methods [20]. Left ventricular ejection fraction was calculated using the 2D-biplane Simpson method.

Prenatal screening with fetal echocardiography was performed according to guidelines and recommendations from the American Society of Echocardiography [21].

This report adhered to the STROBE (Strengthening the Reporting of Observational Studies in Epidemiology) reporting guidelines (see STROBE checklist in the Supplemental Material). The study protocol was approved by the local ethics committee. Written informed consent was obtained from the parents or legal guardians of all patients. The study conformed to the ethical principles outlined of the 1975 Declaration of Helsinki. This study was approved by the Ethics Committee of Bambino Gesù Children’s Hospital, [Protocol No 3565_OPBG_2024].

### Genetic testing

All genetic testing was centralized and performed by the Medical Genetic Laboratories of the Bambino Gesù Children’s Hospital, IRCCS. Genomic DNA was extracted from peripheral blood with Qiagen columns (QIAamp DNA Minikit; Qiagen, Hilden, Germany) according to the manufacturer’s instructions. Next-generation sequencing (NGS) was performed on genomic DNA using the Twist Human Comprehensive Exome Kit (Twist Bioscience) on an Illumina NexSeq550 or NovaSeq6000 platform (Illumina, San Diego, CA, USA), according to the manufacture’s protocol. Variants in genes associated with cardiomyopathies and mitochondrial disease were analyzed and classified according to current ACMG Guidelines [22,23]

### Statistical Analysis

Clinical data are reported using descriptive statistics. Categorical variables are expressed as absolute numbers or percentages, whereas continuous variables were presented as median value and interquartile (IQR) range. Survival analysis and freedom from clinical events, including ICD implantation and cardiac surgery, were assessed using Kaplan Meier curves, and multivariable Cox proportional hazards regression analysis were performed to identify predictors of adverse outcomes. A two-sided *p*-value ≤0.05 was considered statically significant. Patients with missing data for specific clinical outcomes were excluded from analyses involving the corresponding outcome. Statistical analyses were performed using SPSS Statistics 21 (IBM Corporation, Armonk, NY, USA) and R Statistical Software, version 4.5.3 (R Foundation for Statistical Computing, Vienna, Austria).

## RESULTS

### Study Population and Clinical Characteristics at Presentation

Among 321 cases of pediatric patients with HCM referred to and followed in Bambino Gesù Children’s Hospital, 68 (21%) were diagnosed during infancy (<12 months of age). Of these, 51 patients (75%) presented the neonatal period and constituted the study cohort. Prenatal suspicion of HCM was reported in 13 neonates and subsequently confirmed after birth. Five patients (10%) were diagnosed at birth, 24 (47%) between days 1 and 7 of life, and 9 (18%) after the first week (Figure 1). Male sex was slightly predominant (60%), and median body surface area (BSA) at presentation was 0.2 m². Median age at diagnosis was 1 day (IQR 0–6 days), with most patients (82%) presenting within the first week of life.

**Figure 1:**
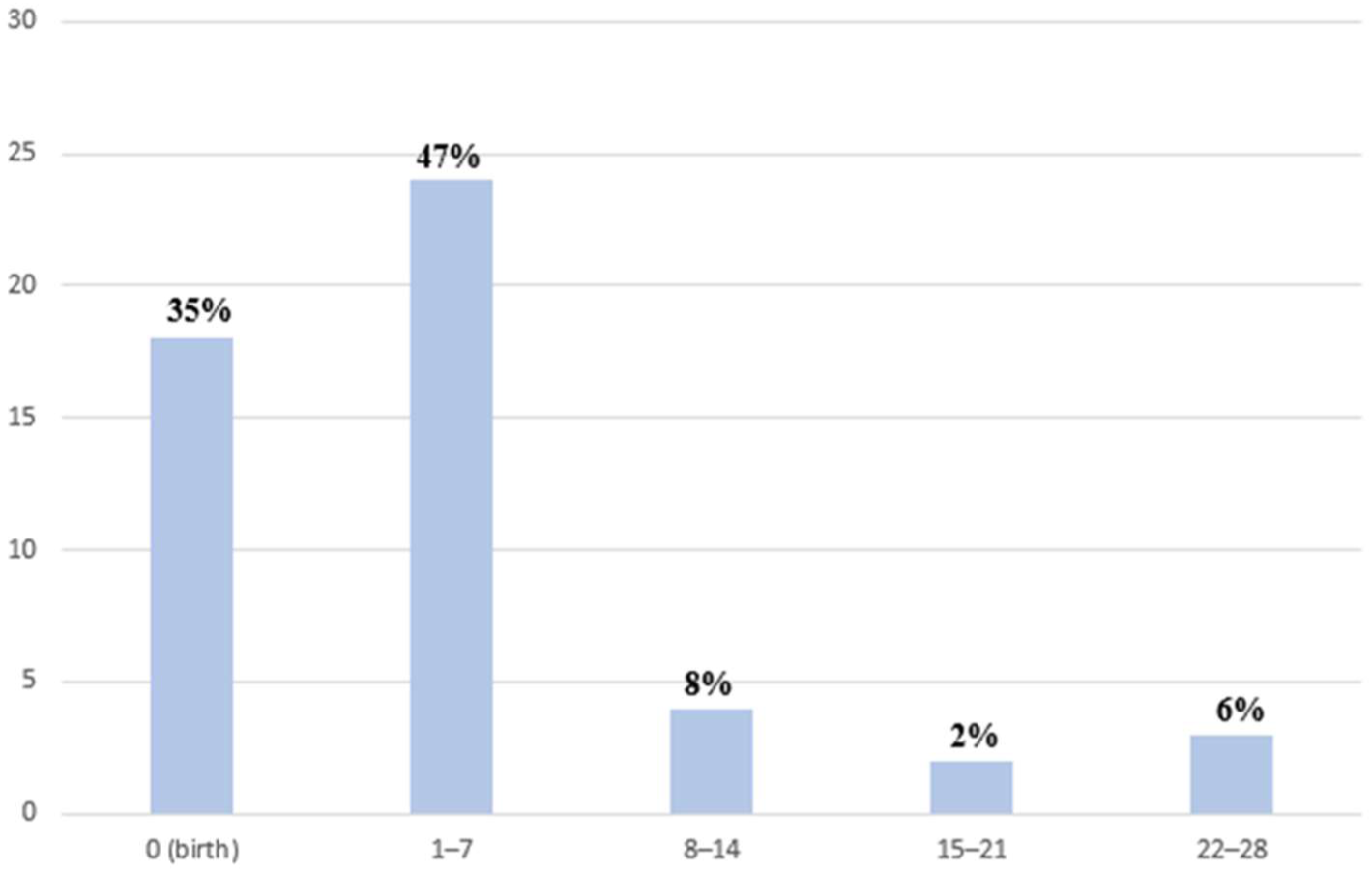
Distribution of age at presentation in neonatal-onset hypertrophic cardiomyopathy. Bar chart showing the distribution of patients according to age at clinical presentation during the neonatal period. The majority of cases were diagnosed during the first week of life, accounting for 47% of the cohort, while 35% were diagnosed at birth or during prenatal evaluation.

### Mode and Timing of Presentation

At initial evaluation, 21 patients (41%) were symptomatic and required hospital admission because of HF manifestations or hemodynamic instability. Clinical presentation included Ross class ≥III symptoms, respiratory distress with tachypnea and increased work of breathing and/or signs of circulatory compromise, corresponding predominantly to stage C HF according to ACC/AHA classification [24]. The prevalence of symptomatic HF at presentation varied according to etiology, occurring in 29% of RASopathies, 25% of those with sarcomeric, 56% of mitochondrial or inborn errors of metabolism (IEM), and 25% of gene-elusive cases (Table 1). The remaining 30 patients (59%) were asymptomatic at presentation. Among the asymptomatic patients: 13 cases (25%) were identified through prenatal screening, 16 patients (31%) were referred after detection of a cardiac murmur during routine clinical evaluation and 1 (2%) additional patient underwent cardiac assessment because of associated dysmorphic features.

**Table 1:** Clinical and echocardiographic characteristics at presentation of neonatal-onset hypertrophic cardiomyopathy, stratified by etiologic group.

|  | *RASopathies*<br>(N=21) | *Sarcomeric*<br>(N=4) | *Mitochondrial*<br>*/ Iem* (N=9) | *Other\**<br>(N=1) | *Gene Elusive*<br>(N=16) |
| --- | --- | --- | --- | --- | --- |
| **Demographic data** |  |  |  |  |  |
| *Age (days)* | 1 (0–7) | 4 (0–4) | 3 (0–8) | 0 | 4 (0–10) |
| *Male sex* | 11 (52%) | 2 (50%) | 6 (67%) | 1 (100%) | 8 (50%) |
| *BSA* | 0.21 (0.2–0.23) | 0.0 (0.2–0.21) | 0.2 (0.18–0.22) | 0.21 | 0.21 (0.2–0.22) |
| **Clinical data** |  |  |  |  |  |
| *Symptomatic* | 6 (29%) | 1 (25%) | 5 (56%) | 1 (100%) | 4 (25%) |
| *Ross Class I* | 4 (19%) | 2 (50%) | 2 (22%) | 0 | 4 (25%) |
| *Ross Class II* | 5 (24%) | 0 | 2 (22%) | 0 | 5 (31%) |
| *Ross Class ≥ III* | 5 (24%) | 1 (25%) | 2 (22%) | 0 | 5 (31%) |
| *Cardiac shock* | 7 (33%) | 1 (25%) | 3 (33%) | 1 (100%) | 2 (13%) |
| **Echocardiogram** |  |  |  |  |  |
| *LVEF (%)* | 63 (60–70) | 62 (59–64) | 70 (65–72) | 68 | 65 (60–70) |
| *LPWT (mm)* | 5.0 (4–6) | 5.5 (4–6.5) | 4.5 (4–6) | 8 | 5 (3–6.5) |
| *IVS (mm)* | 9 (6–12) | 10 (9–13) | 7 (6–10) | 14 | 8 (6–12) |
| *IVS Z-Score* | 5 (2–7) | 3 (2–4) | 3.5 (2–6) | 5.5 | 4 (2–6) |
| ***LVOT-O (n)*** | ***6 (29%)*** | ***2 (50%)*** | ***1 (11%)*** | ***1 (100%)*** | ***3 (19%)*** |
| ***LVOT-O*** | ***55 (36–80)*** | ***80 (75–85)*** | ***45 (40–60)*** | ***100*** | ***60 (35–100)*** |
| ***RVOT-O (n)*** | ***5 (24%)*** | ***0*** | ***2 (22%)*** | ***1 (100%)*** | ***2 (13%)*** |
| ***RVOT-O*** | ***60 (40–75)*** | ***0*** | ***40 (40–60)*** | ***80*** | ***60 (60–60)*** |
| ***MR ≥ moderate*** | ***8 (38%)*** | ***2 (50%)*** | ***3 (33%)*** | ***0*** | ***2 (12.5%)*** |

***Other\*:*** one patient with 3p11.1 microduplication
***Legend:*** Age = at onset; BSA = body mass index; LVOT-O = Left ventricular outflow tract obstruction (mmHg); RVOT-O = right ventricular outflow tract obstruction (mmHg); LPWT = Left Posterior Wall Thickness (mm); IVS = interventricular septum (mm); LVEF = Left ventricular ejection fraction (%); MR= mitral regurgitation.

Clinical presentation differed according to age at diagnosis. Patients identified at birth were predominantly asymptomatic (72%), largely due to prenatal screening and planned delivery with immediate transfer to the neonatal intensive care unit. Conversely, five neonates (28%) were symptomatic on the first day of life, typically presented with severe respiratory distress and/or oxygen desaturation. HF manifestations became more common during the first week of life (50%) and reached the highest prevalence during the second week (75%). At this age, symptoms were mainly characterized by poor feeding and failure to thrive. Beyond the second week, most patients remained asymptomatic; symptomatic patients mainly showed sweating during feeding, tachypnea and growth impairment (Figure 2). In these later-presenting cases, referral was generally prompted by family screening or detection of a cardiac murmur.

**Figure 2:**
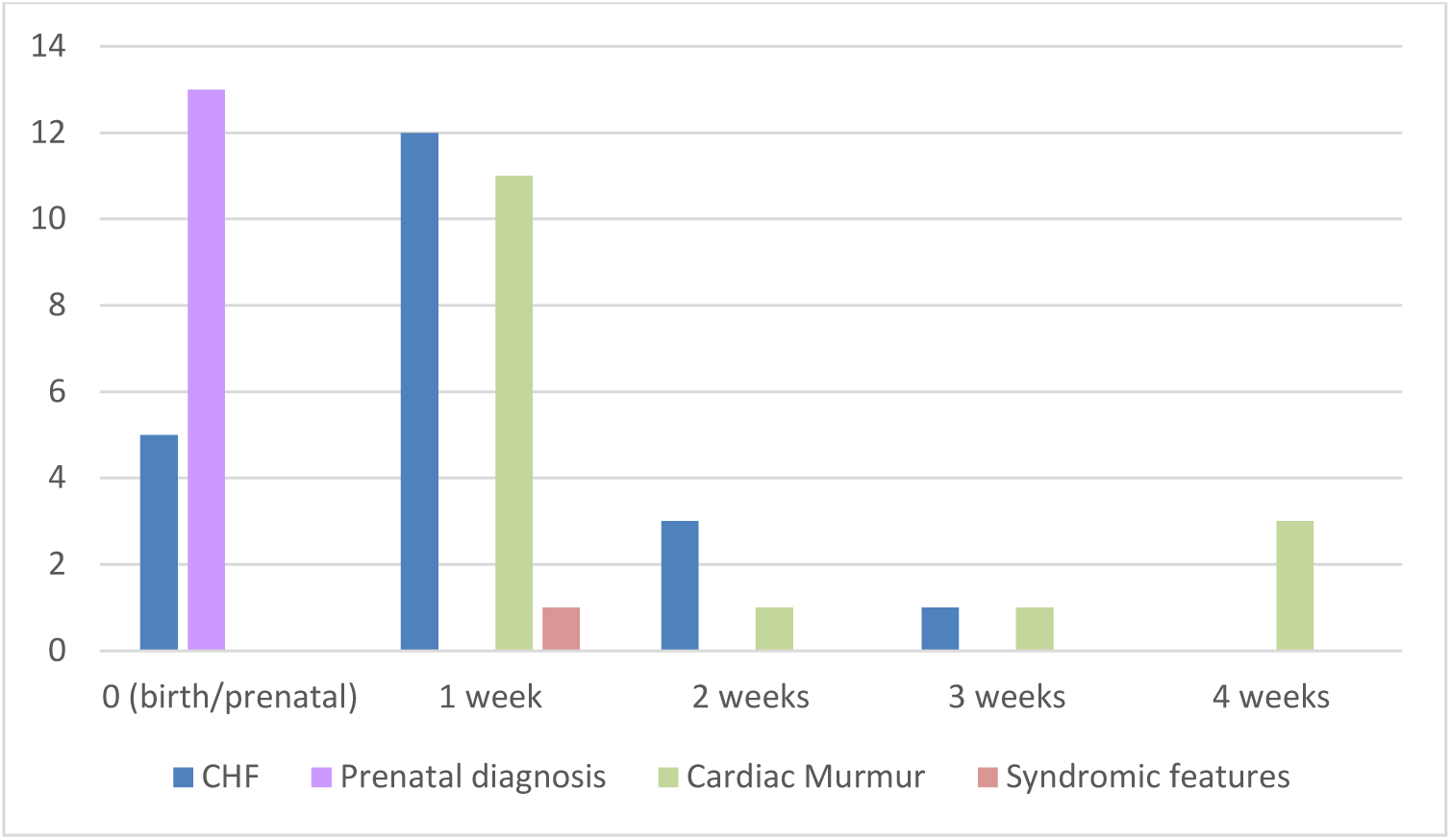
Mode of presentation according to age at diagnosis. Stacked column chart illustrating the distribution of different modes of presentation across age groups. Symptomatic patients presented with congestive heart failure (CHF), while asymptomatic cases were identified through prenatal diagnosis, detection of a cardiac murmur, or syndromic features. The figure highlights the predominance of prenatal diagnosis at birth and the increasing contribution of cardiac murmurs to diagnosis in later time points.

Medical therapy was initiated in symptomatic patients and consisted primarily of beta-blockers treatment, most commonly propranolol, progressively uptitrated to the maximum tolerated or clinically effective dose. During FU, 41% of patients required high dosage of beta blockers (≥ 6mg/kg/day) [25–26]. Ultimately, all patients developed symptoms during the disease course and received pharmacological treatment.

### Etiologic Distribution of Neonatal HCM

Among the 51 neonates with HCM, RASopathies represented the most common etiologic category, affecting 21 patients (41%). Gene-elusive HCM accounted for 16 cases (31%), followed by mitochondrial or IEM- related disease in 9 patients (18%), sarcomeric HCM in 4 (8%), and other genetic conditions in 1 patient (2%) (Figure 3).

**Figure 3:**
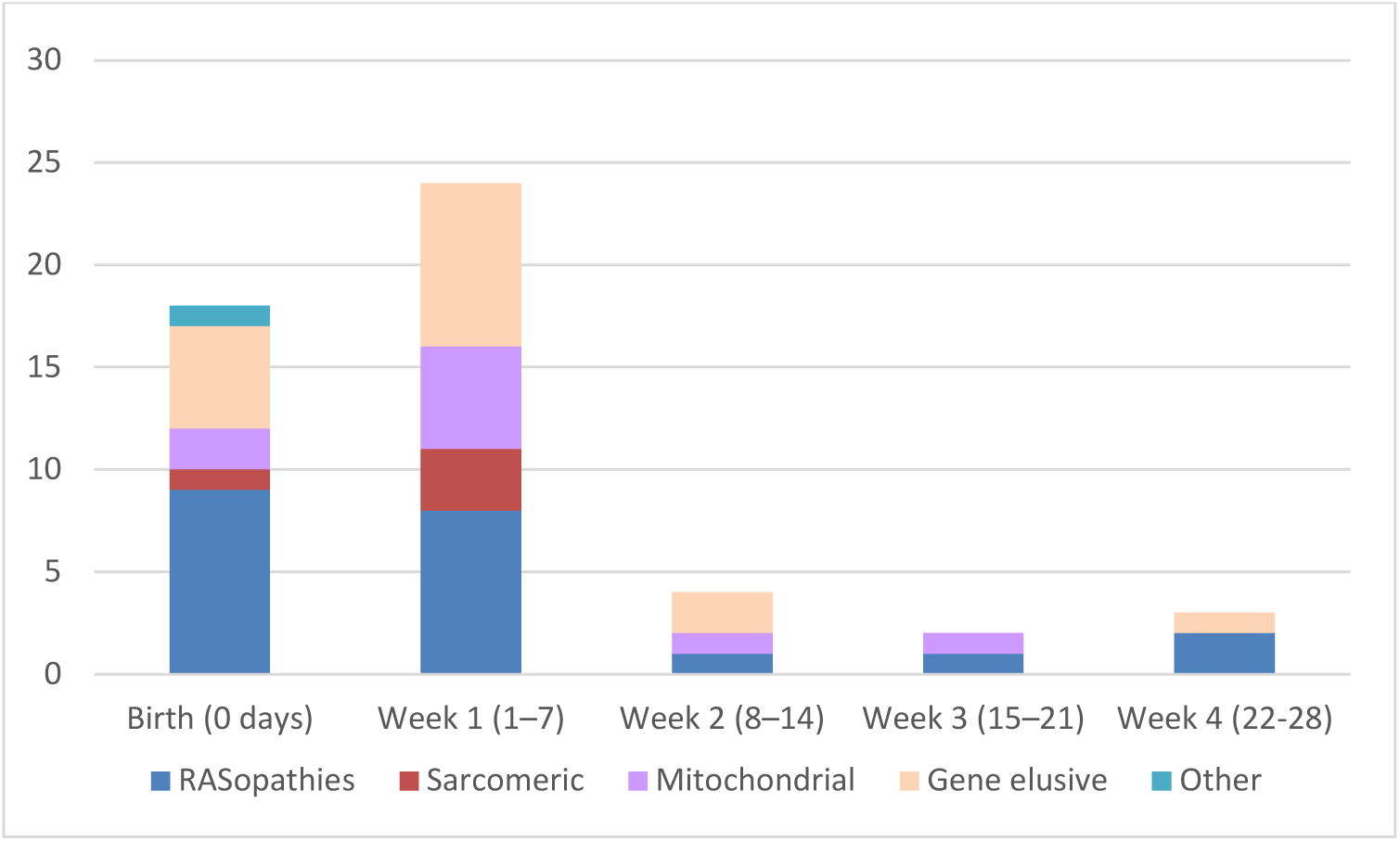
Distribution of neonatal-onset hypertrophic cardiomyopathy according to week of presentation and genetic etiology. Stacked bar chart showing the distribution of the 51 patients included in the cohort according to week of clinical presentation and underlying etiologic group. Most patients presented within the first week of life, accounting for the largest proportion of cases. RASopathies represented the most frequent etiologic category and were distributed throughout the neonatal period, while sarcomeric forms were observed exclusively during the first week of life. Mitochondrial and gene-elusive cases were also predominantly diagnosed early in life.

Within the RASopathy subgroup, variants in *PTPN11* were the most frequently identified (9/21, 43%), followed by *SOS1*, *RAF1*, and *LZTR1* variants (each 3/21, 14%), *RIT1* (2/21, 10%), and *PPP1CB* (1/21, 5.0%). Sarcomeric HCM was associated with variants in MYH7 (2/4, 50%), *MYBPC3* (1/4, 25%), and *MYL2* (1/4, 25%). Patients classified as gene-elusive showed no pathogenic or likely pathogenic variants in currently recognized HCM- associated genes, whereas the “other genetic conditions” category included one patient carrying a 3p11.1 microduplication (Table 2).

**Table 2.**
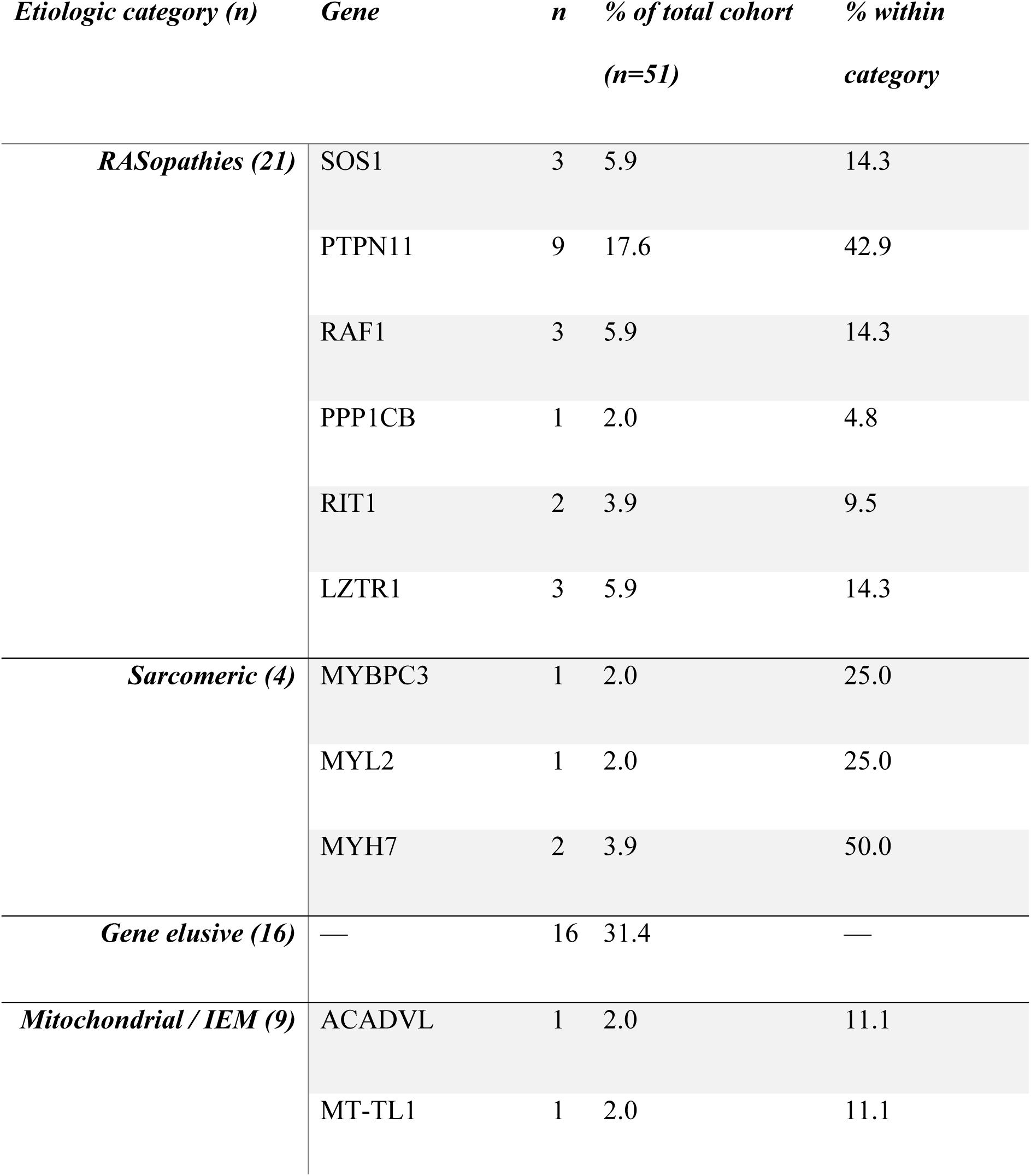
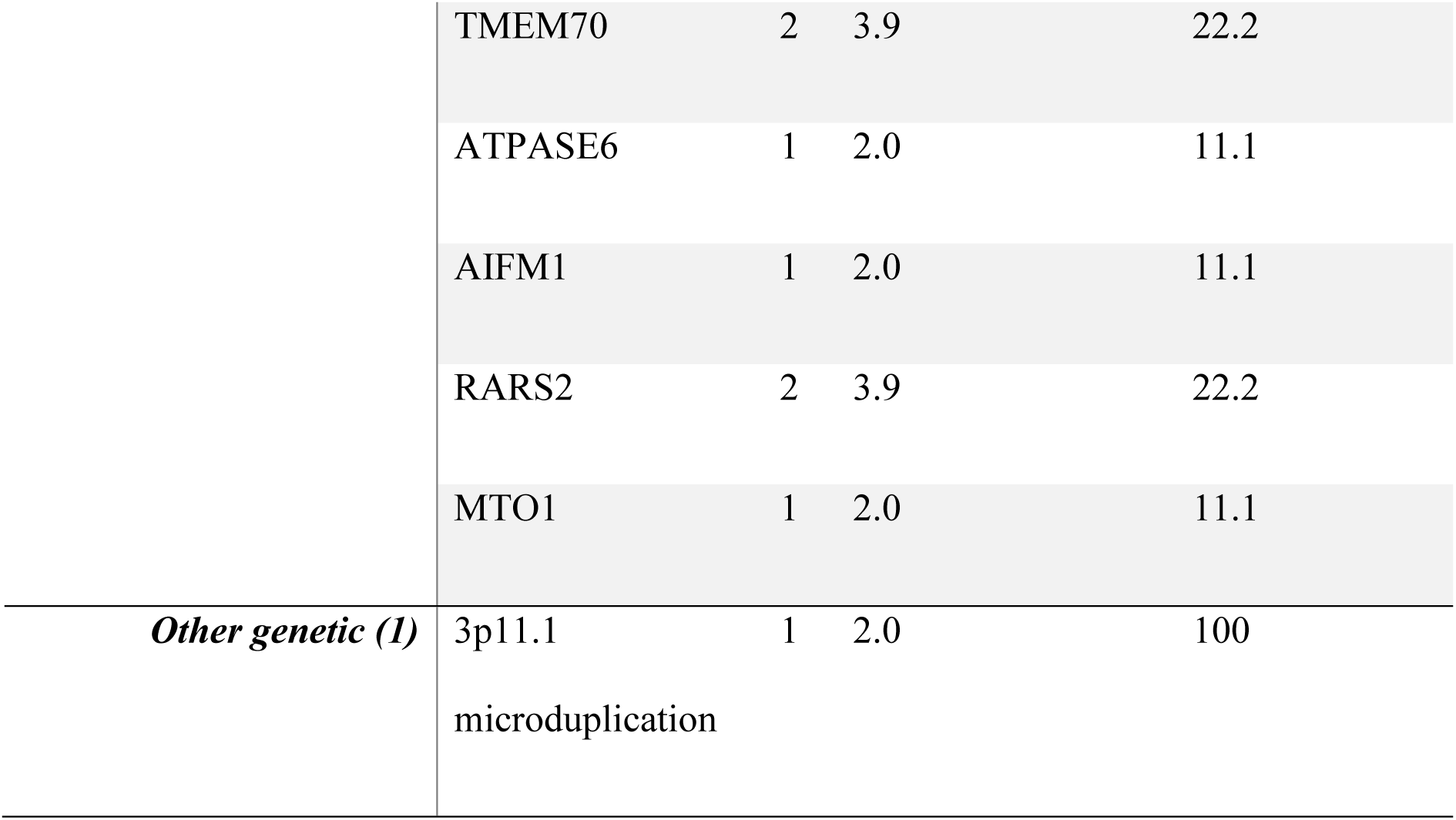
Genetic and etiologic distribution of neonatal-onset hypertrophic cardiomyopathy. Distribution of patients according to major etiologic categories and specific gene variants. Percentages are shown for the total cohort and within each category. Gene-elusive patients had no pathogenic variant identified in known HCM-associated genes.

RASopathies-associated HCM was distributed throughout the neonatal period, although most cases presented during the first week of life (17/21, 81%). Sarcomeric forms were exclusively diagnosed within the first week of life, while mitochondrial/IEM-related and gene-elusive forms also predominantly manifested early after birth. Presentations beyond the second week were uncommon and mainly involving RASopathy-associated or and gene-elusive disease (Figure 3). LVOT obstruction was observed in up to 50% of sarcomeric cases and in 29% of RASopathies. Moderate or greater mitral regurgitation was present in 38% of RASopathy patients and 50% of sarcomeric cases. (Table 1).

### Clinical Outcomes

#### Short-Term Outcome

In the neonatal-onset cohort, overall survival at 2 years was 92% (CI 95% 85-99.5), and cardiac death was 96% (CI 95% 90.8-100), compared with 100% in patients diagnosed after the neonatal period >28^th^ day but <1^st^ year of life (log-rank=0.2; Figure 4 A-B).

**Figure 4:**
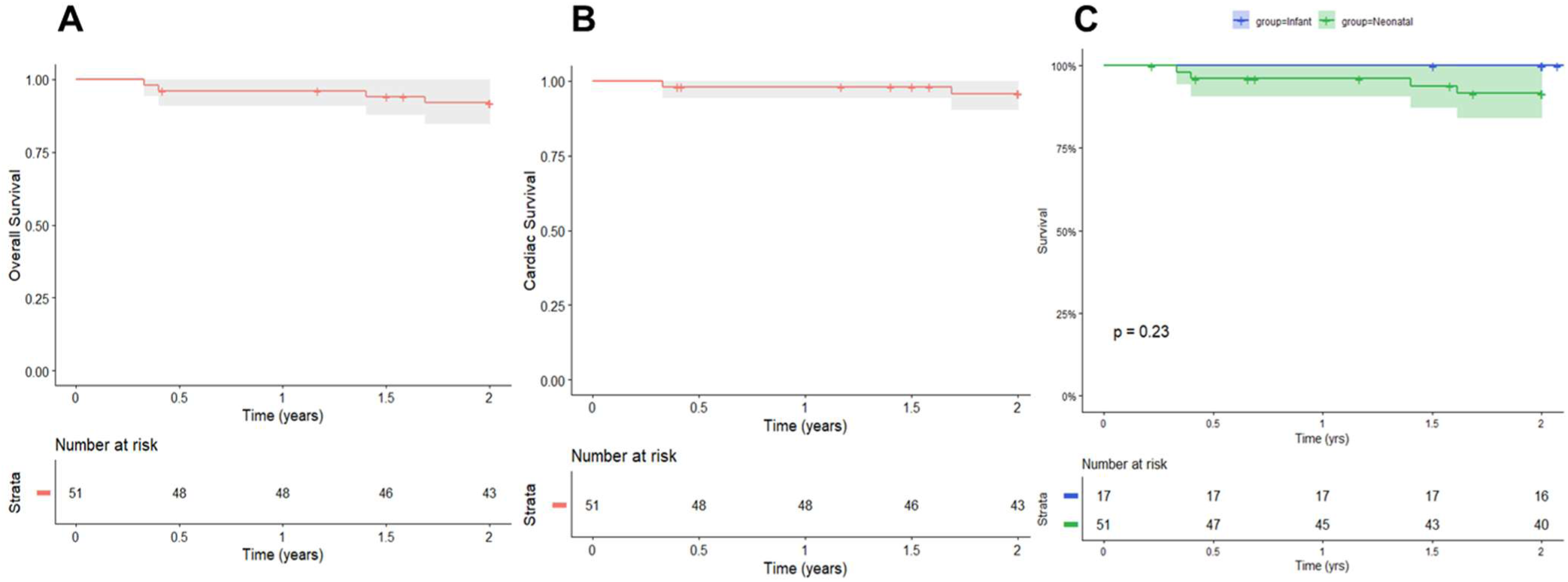
A) Overall neonatal survival at 2 years; B) Cardiac neonatal survival; C) Overall survival comparing neonates with infants. (A) Kaplan–Meier estimate of overall survival in the neonatal-onset cohort during the first 2 years of FU. (B) Kaplan–Meier estimate of cardiac survival in the neonatal-onset cohort during the first 2 years of FU. (C) Kaplan–Meier comparison of overall survival between neonatal-onset and infantile-onset hypertrophic cardiomyopathy cohorts.

During FU, one patient died because of septic shock, and one patient underwent heart transplantation. One died suddenly at home. One additional patient experienced resuscitated sudden cardiac arrest.

Within the first two years of life, six patients (12%) required cardiac surgery, including septal myectomy and right ventricular outflow tract (RVOT) obstruction relief procedures; one patient also underwent surgical correction of a mitral cleft. One death occurred for untractable postoperative infection. No ventricular assist device implantation was recorded.

Cardiac events occurred predominantly in symptomatic patients and were mainly clustered among neonates presenting at birth or within the first two weeks of life.

#### Long-Term Outcome

Median FU duration was 6.9 years (IQR 3.6–12.0, range 0.3-32.7 years), with complete FU available for all patients.

Long-term overall survival in the neonatal-onset cohort was 88% (CI 95% 70-96) at 30 years, whereas no deaths occurred in the infantile-onset group (log-rank=0.094; Figure 5). Cardiac survival in the neonatal-onset cohort was 92% (CI 95% 88.5-99-7) at 30 years. After 2 years, during long term FU, one patient underwent heart transplantation.

**Figure 5:**
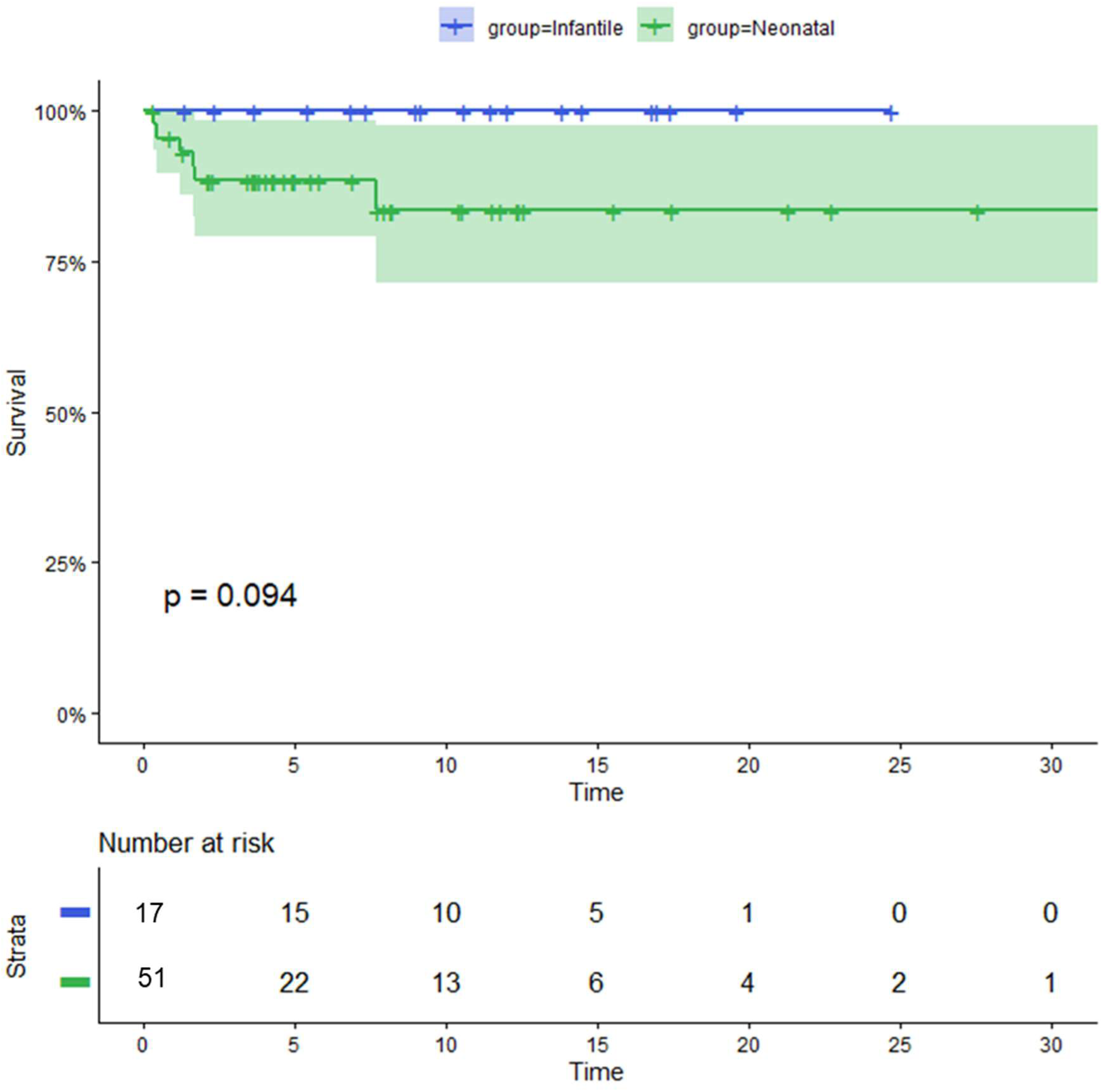
Long-term survival of neonates and infantile-form. Kaplan–Meier analysis comparing long-term survival between neonatal-onset and infantile-onset hypertrophic cardiomyopathy cohorts. Overall survival at 30 years was 88% in the neonatal-onset cohort, whereas no deaths occurred in the infantile-onset group (log-rank P=0.094).

ICD were implanted in four patients (8%): three for primary prevention and one for secondary prevention following resuscitated sudden cardiac arrest. One patient with ICD in primary prevention experienced an appropriate ICD shock for ventricular fibrillation during FU. Survival free from ICD implantation was 40% (CI 95% 27-55) at 15 year (Figure 6A). ICD implantation was significantly different at 15 years according to etiology, more pronounced in those affected by sarcomeric or gene elusive and syndromic variants (log rank= 0.03, fig 6B). Overall, 11 patients (21%) underwent cardiac surgical procedures, including septal myectomy and RVOT interventions. Survival free from myectomy was 47% (CI 95% 37-60) at 15 years (Figure 7A). According to etiology, myectomy has a meaningful impact in those with syndromic variants (log-rank = 0.01, fig 7B). At 30 years, no significant difference was detected for ICD and myectomy.

**Figure 6:**
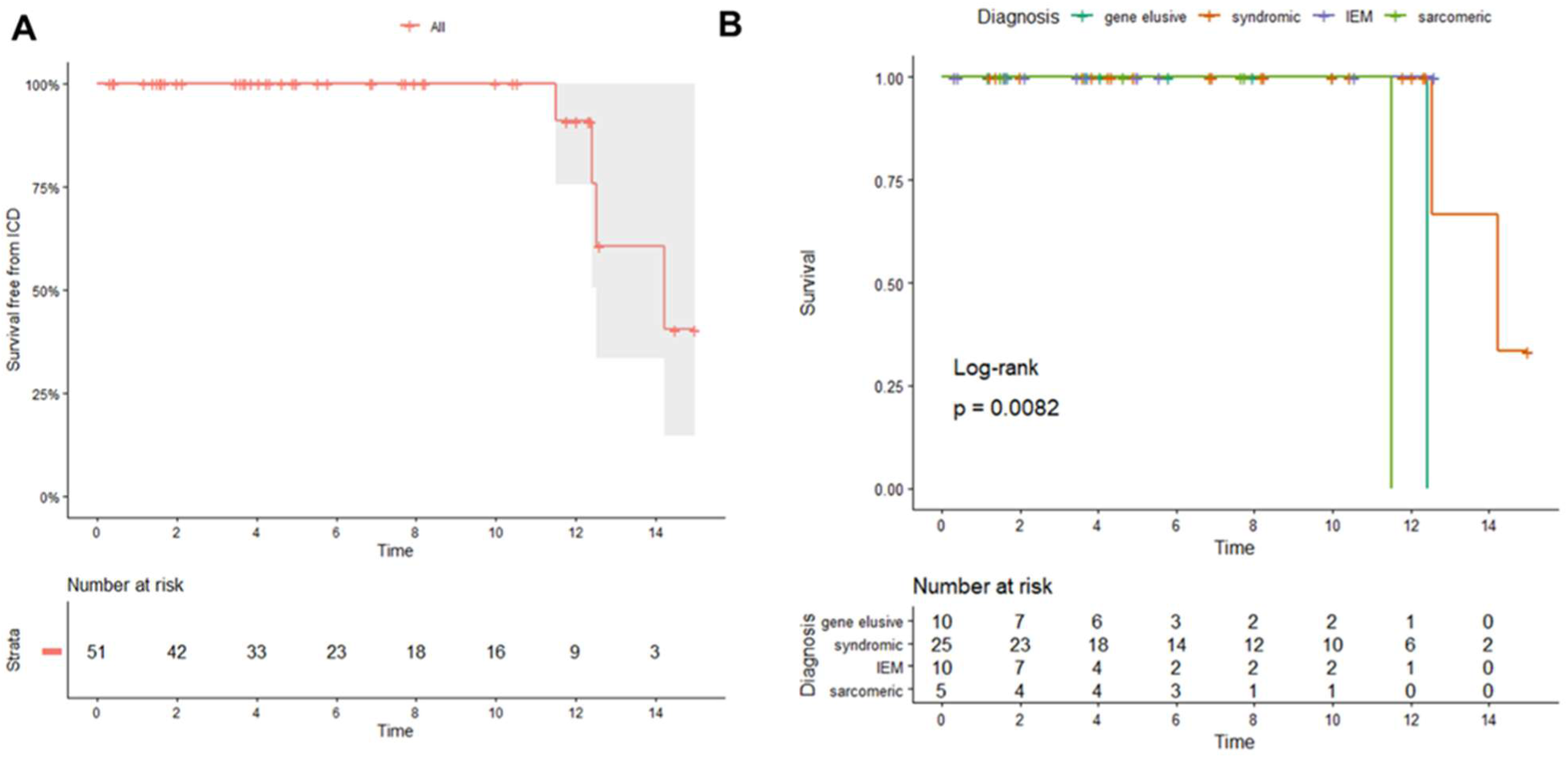
A) Overall long-term survival free from ICD implantation. B) Long-term survival free from ICD implantation according to etiology. (A) Kaplan–Meier estimate of survival free from ICD implantation during FU, at 15 years. (B) Kaplan–Meier estimate of survival free from ICD implantation during FU, at 15 years, according to etiology.

**Figure 7:**
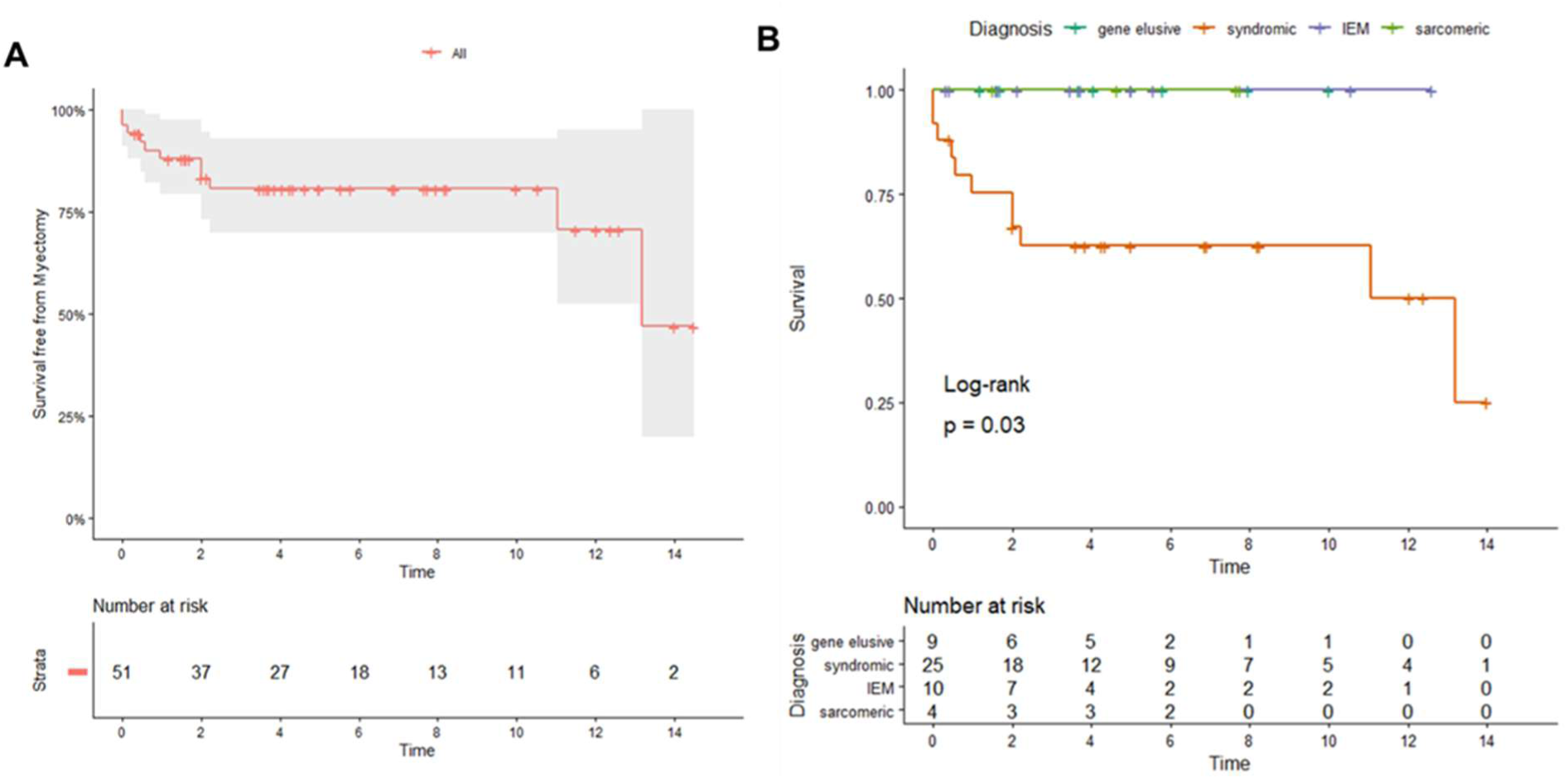
A) Overall long-term survival free from myectomy. B) Long-term survival free from myectomy according to etiology. (A) Kaplan–Meier estimate of survival free from myectomy during FU, at 15 years. (B) Kaplan–Meier estimate of survival free from myectomy during FU, at 15 years, according to etiology.

Major adverse cardiac events and cardiac death occurred exclusively in patients with very early disease onset, defined as presentation within the first 14 days of life (median age 1 day [IQR 0–4]). All deaths and heart transplantations occurred in neonates presenting within the first three weeks of life, and ICD implantation was similarly confined to this subgroup. The only episode of resuscitated sudden cardiac arrest occurred in a patient diagnosed at birth. Cox regression analysis did not identify a statistically significant predictor factor for MAEs or cardiac death, likely because of the limited sample size.

## DISCUSSION

This study provides one of the largest dedicated analyses of neonatal-onset HCM and highlights the distinctive clinical and prognostic profile of HCM presenting during the first weeks of life. Several relevant findings emerge from our cohort.

First, neonatal HCM represented a substantial proportion of infantile HCM cases referred to a tertiary pediatric cardiomyopathy center, emphasizing that presentation during the neonatal period is not exceptional in contemporary pediatric practice. Data focusing specifically on neonatal presentations are limited, as most studies focus on infantile or broader pediatric population [2–4, 27, 28]. Previous studies [1–4] have consistently shown that age at diagnosis is a major determinant of prognosis, with patients presenting within the first year of life, especially within earliest months of life, experiencing significantly higher mortality and transplantation rates compared with older children. Importantly, in our cohort most patients were diagnosed within the first week of life, frequently through prenatal suspicion or early postnatal evaluation prompted by respiratory distress or cardiac murmurs. These findings underline the increasing contribution of prenatal and early neonatal cardiovascular screening in identifying severe cardiomyopathic phenotypes before overt clinical deterioration.

Second, our data confirm the marked etiologic heterogeneity of neonatal HCM, but also demonstrate the predominance of syndromic and non-sarcomeric etiologies in this age group. RASopathies accounted for more than 40% of cases and represented the most frequent identifiable cause, whereas classic sarcomeric HCM was relatively uncommon [11,27]. This distribution differs substantially from later childhood and adult HCM, where sarcomeric disease predominates, and reinforces the concept that neonatal HCM should be considered a biologically distinct entity. The high prevalence of gene-elusive patients also reflects the persistent diagnostic challenges in neonatal cardiomyopathy despite contemporary next-generation sequencing strategies.

Third, the mode and timing of presentation strongly influenced disease severity and long-term outcomes. Patients presenting within the first days of life were more likely to develop severe HF, require intensive care support, and experience adverse cardiac events. Notably, all deaths, heart transplantations, and malignant arrhythmic events occurred exclusively among patients presenting within the first weeks of life. These findings suggest that very early clinical onset may represent a surrogate marker of aggressive myocardial disease biology rather than simply an earlier stage of the same disease spectrum.

Although in our cohort overall survival was relatively favorable considering the severity of presentation, neonatal-onset HCM remained associated with substantial long-term morbidity, especially in syndromic patients. In previous studies [1–4] the disease burden of infantile HCM was characterized by a high risk of mortality, particularly in those with symptoms and syndromic etiology [6]. In our cohort, a considerable proportion of patients required high-dose beta-blocker therapy [29–34], surgical relief of ventricular outflow tract obstruction, or ICD implantation during FU. The need for septal reduction procedures and the occurrence of arrhythmic complications indicate that neonatal HCM may evolve into a chronic and progressive cardiomyopathic phenotype requiring lifelong specialized management. The difference detected after 15 years underline the importance of adolescence turning point, requiring a specific follow up. During adulthood, these differences based on etiology resulted attenuated.

Interestingly, symptomatic HF at presentation was more frequent among mitochondrial and metabolic disorders, whereas obstructive physiology was particularly represented in sarcomeric and RASopathy-associated forms. These observations further support the notion that genotype substantially influences early phenotype expression and clinical trajectory in neonatal HCM. However, despite clear phenotypic trends, multivariable analysis did not identify independent predictors of adverse outcome, likely because of the relatively limited sample size and low number of events.

Our findings also have important implications for prenatal counseling and postnatal management. The increasing recognition of fetal myocardial hypertrophy raises difficult prognostic and therapeutic questions, in particular in the absence of a defined molecular diagnosis. In this setting, the present study provides clinically relevant information showing that outcomes are heterogeneous and not universally poor, although patients presenting within the first two weeks of life represent a remarkably high-risk subgroup warranting close monitoring and early referral to expert cardiomyopathy centers.

In conclusion, neonatal-onset HCM represents a rare but highly heterogeneous condition characterized by early presentation, predominance of syndromic and non-sarcomeric etiologies, and significant long-term cardiovascular morbidity. Presentation within the first days of life identifies a subgroup at particularly high risk for HF progression and major adverse cardiac outcomes. Early recognition, comprehensive genetic evaluation, and long-term multidisciplinary FU are essential to optimize management and improve outcomes in this vulnerable population.

## LIMITATIONS OF THE STUDY

The study should be interpreted in light of several limitations. Its retrospective design and the long enrollment period may have introduced heterogeneity in diagnostic approaches, genetic testing availability, and management strategies over time. In addition, the study was conducted in tertiary referral centers, potentially leading to referral bias toward more severe phenotypes. Finally, despite representing one of the largest neonatal HCM cohorts reported to date, the relatively small number of adverse events limited statistical power for risk stratification analyses.

## Data Availability

The data that support the findings of this study are available from the corresponding author upon reasonable request.

## Acknowledgments

Our Centre is a health care provider of the European Reference Network (ERN) GUARD-Heart. The authors gratefully acknowledge all patients and families included in this study, as well as the physicians and healthcare professionals from the participating centers who contributed to clinical data collection and multidisciplinary care.

## APPENDIX

### Cooperating Investigators†

University “La Sapienza”, Rome, Italy, *Prof. Paolo Versacci*

Fondazione Policlinico Universitario “Agostino Gemelli” IRCCS, Rome, Italy, *Dr Luciano Pasquini, Dr Angelica Delogu*

Antonio Perrino Hospital, Brindisi, Italy, *Dr. Enrico Rosati*.

Azienda Ospedaliera Universitaria Ospedali Riuniti di Ancona Umberto I G M Lancisi G Salesi, Ancona, Italy, *Dr. Emanuela Berton*.

---

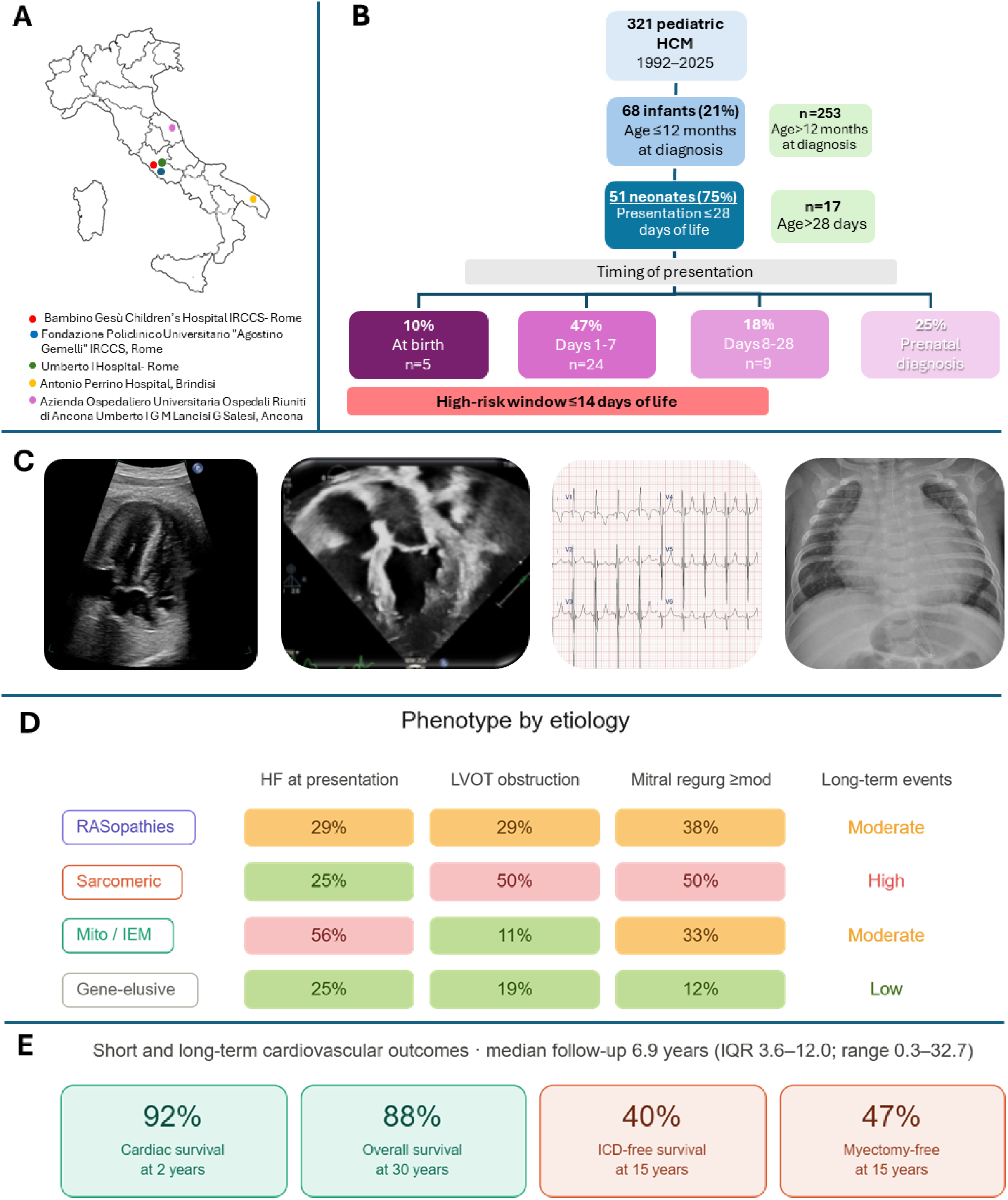

